# Functional status, mood state, and physical activity among women with post-acute COVID-19 syndrome

**DOI:** 10.1101/2022.01.11.22269088

**Authors:** Stephen J. Carter, Marissa N. Baranauskas, John S. Raglin, Bernice A. Pescosolido, Brea L. Perry

**Author notes:** The present work was funded, in part, by the Indiana Clinical and Translational Sciences Institute by grant number UL1TR002529 from the NIH National Center for Advancing Translational Sciences, Clinical and Translational Sciences Award. Corresponding Author: Stephen J. Carter, M.S., Ph.D., Assistant Professor, Department of Kinesiology, School of Public Health – Bloomington, Indiana University, 1025 E 7^th^ Street Suite 044.

## Abstract

**Objectives:** While organ-specific pathophysiology has been well-described in SARS-CoV-2 infection, less is known about the attendant effects on functional status, mood state and leisure-time physical activity (PA) in post-acute COVID-19 syndrome.

**Methods:** A case-control design was employed to recruit 32 women (*n* = 17 SARS-CoV-2; *n* = 15 controls) matched on age (54 ± 12 years), body mass index (27 ± 6 kg/m^2^), smoking status, and history of cardiopulmonary disease. Participants completed a series of assessments including the Modified Pulmonary Functional Status and Dyspnea Questionnaire (PFSDQ-M), Profile of Mood States (POMS), and Godin-Shephard Leisure-Time PA.

**Results:** SARS-CoV-2 participants exhibited poorer functional status (*p* = 0.008) and reduced leisure-time PA (*p* = 0.004) compared to controls. Significant between-group differences were also detected for the POMS total mood disturbance with sub-scale analyses revealing elevated tension, confusion, and lower vigor among SARS-CoV-2 participants (all *p*-values < 0.05). The number of SARS-CoV-2 symptoms (e.g., loss of taste / smell, muscle aches etc.) were associated (*r* = 0.620, *p* = 0.008) with confusion.

**Conclusion:** The sequela of persistent SARS-CoV-2 symptoms elicit clear disturbances in functional status, mood state, and leisure-time PA among women with post-acute COVID-19 syndrome.

## INTRODUCTION

Worldwide estimates indicate over 300 million individuals have been diagnosed with severe acute respiratory syndrome coronavirus 2 (SARS-CoV-2), with the United States accounting for approximately 20% of total cases [1]. Though vaccination efforts have reduced mortality incidence, emerging hot spots and SAR-CoV-2 variants remain a significant threat to personal and public health [2]. Recent data reveals some individuals experience a delayed recovery with symptoms persisting three to four weeks (or longer) beyond initial SARS-CoV-2 diagnosis – a condition now recognized as post-acute COVID-19 syndrome [3]. Among these individuals, latent effects vary considerably but generally include dyspnea, fatigue, mental fog and/or sensory disturbances. Understandably, such complications bear clinical and practical relevance for health and wellness as the number of individuals experiencing lingering SARS-CoV-2-related symptoms grows. While studies have widely characterized clinical outcomes among individuals hospitalized with severe-to-critical SARS-CoV-2 infection – most recover from the acute effects of the virus. Interestingly, illness severity following primary infection, does not appear to be associated with the possibility of longer-term SARS-CoV-2 symptoms [4].

Recent data from the Mayo Clinic’s COVID-19 Activity Rehabilitation Program indicate women outnumber men 3 to 1 in pursuit of treatment for post-acute COVID-19 syndrome [5]. In France, a similar proportion (sex ratio 4:1) has been reported in women approximately 40 years of age seeking treatment for longer-term SARS-CoV-2 symptoms [6]. This, in turn, raises important questions about the presentation and functional consequences of post-acute COVID-19 syndrome in women – especially since women tend to exhibit greater rates of age-related disability than men [7].

A regrettable consequence of the on-going COVID-19 pandemic involves extensive disruption in daily routines affecting work and leisure-time activities [8]. Government lockdowns have restricted public life such that, compared to pre-restriction levels, self-reported physical activity (PA) was reduced by 41% in a multi-national cohort [9]. It is currently unknown whether some of the reduction in PA may be attributable to longer-term effects of SARS-CoV-2 among infected persons. Owing to the systemic psychological benefits of leisure-time PA on variables such as mood state, declining PA may have a negative impact on psychological well-being, including anxiety and depression. Smith and colleagues have shown poorer mental health during the COVID-19 pandemic correlates with being female, younger age, a lower annual income, and having multiple comorbidities [10]. Despite the extensive literature describing the serious physical effects of the SARS-CoV-2 virus in vulnerable populations, little is known about possible relationships between functional status, mood state, and PA behavior in post-acute COVID-19 syndrome.

Undoubtedly, the progressive burden of SARS-CoV-2 warrants further investigation into the consequences of post-acute COVID-19 syndrome. Therefore, the present work compared functional status, mood state, and leisure-time PA in women with and without a history of SARS-CoV-2 matched for age and body mass index (BMI). We hypothesized that SARS-CoV-2 participants would have: 1) poorer functional status, 2) greater mood disturbance, and 3) report less leisure-time PA compared to women without a history of SARS-CoV-2. Such information may inform targeted interventions designed to attenuate the burdens of post-acute COVID-19 syndrome in women.

## METHODS

### Participants

Women *at least* four weeks to four months from having a positive laboratory test for SARS-CoV-2 and women without a history of SARS-CoV-2 (i.e., controls) were enrolled in a case-control design. Eligible participants recruited from the community within a 115-mile radius of Bloomington, Indiana were matched on age and BMI. Exclusion criteria were individuals < 18 or > 75 years of age, BMI < 18.5 kg/m^2^ or > 50 kg/m^2^, currently pregnant, lactating or trying to become pregnant, documented history of pulmonary disease and/or self-reported use of tobacco products within the previous six months. SARS-CoV-2 participants met the criteria for mild-to-moderate illness severity as defined by the National Institutes of Health [11]. Consistent with the working definition of post-acute COVID-19 syndrome reported by Datta et al. [12] and Greenhalgh et al. [13], all SARS-CoV-2 participants (apart from one) reported symptoms beyond 3-4 weeks from initial illness onset. The single (*n* = 1) SARS-CoV-2 participant that did not meet the definition of post-acute COVID-19 syndrome reported a loss of taste and smell lasting 20 days, and as such, were included in statistical analyses. For exploratory purposes, sub-analyses were performed by dichotomizing the SARS-CoV-2 participants by those who were reporting symptoms at the time of testing (i.e., symptomatic) versus those who were not (i.e., asymptomatic). Study procedures were performed in accordance with the ethical guidelines set forth by the Declaration of Helsinki and local institutional review board (IRB-2004439367). Written informed consent was obtained from all participants before study involvement.

### Procedures and Instruments

After an initial phone screen, prospective participants were invited for an onsite visit to the Indiana University Human Performance Laboratory. Participants were measured for: standing height (stadiometer), body mass, body composition, resting heart rate (HR), and resting blood pressure. BMI was determined by dividing body mass in kilograms by standing height in meters squared. Body composition was measured via multi-frequency bioelectrical impedance (MC-780U, Tanita Corporation, Tokyo, Japan) to the nearest 0.5%. Heart rate and blood pressure were taken from the brachial artery via automated monitoring system (CT40, SunTech Medical, Morrisville, SC). Rate-pressure product (RPP), which provides a non-invasive index of myocardial workload, was calculated by dividing the product of HR and systolic blood pressure by 100 [10]. A questionnaire was administered to obtain descriptive information including age (verified by driver’s license), education, employment, marital status, yearly income, and current medications. A detailed inventory of SARS-CoV-2 symptoms were also collected.

The Profile of Mood States (POMS) was used to assess both specific and general mood. The POMS is a 65 item Likert format (0 = *“not at all”* and “4 = *extremely)* questionnaire that measures the specific mood factors of tension, depression, anger, vigor, fatigue and confusion [14]. A widely used higher order measure of total mood disturbance (TMD) was assessed by adding the negative mood variables and subtracting the positive variable of vigor. Participants completed the POMS on the basis of the standard instructional format (i.e., *“last week including today”*), which yields scores of moderate stability known to be responsive to major interventions including longer-term participation on PA programs. Of note, in situations where researchers are interested in acute mood state responses to an exposure of a single stimulus (e.g., exercise), the instructional set is altered (i.e., *“how do you feel right now”*) [15]. The norms provided in the POMS are not closely comparable to the present sample, and as such, results from Nyenhuis et al. [16] that assessed the POMS in 400 adults (52% women, age = 44 ± 18 y) were used to standardize scores to a population norm (Z-score = observed value – population mean / population SD). Z-scores were converted to T-scores with a mean of 50 and SD of 10 [T-score = 50 + (10 x Z-score)].

A modified Pulmonary Functional Status and Dyspnea Questionnaire (PFSDQ-M) [17] was used to examine functional status at the time of testing. Participants were asked to rate the degree of shortness of breath and fatigue they were experiencing during 10 activities of independent living (e.g., showering, putting on a shirt, preparing a snack, climbing 3 stairs, walking on uneven ground etc.). The Likert-scales were anchored at *“0 = no shortness of breath / fatigue”* and *“10 = very severe shortness of breath / fatigue*.*”* The final section questioned participants about how their involvement in each activity had changed compared to pre-SARS-CoV-2 diagnosis (or over the past 6-months among controls). The scale was anchored at *“0 = no change” and “10 = can no longer perform*.*”* A total score (ranging from 0 to 300) was determined by the summation of all components with a higher score indicating poorer functional status.

An eight-item scale was used to measure walking self-efficacy in which participants were asked to rate their level of confidence (0%-100% in 10% increments) in ability to walk at a moderately-fast pace without stopping for 5 min up to 40 min in 5-min increments [18]. Of note, walking self-efficacy is significantly associated with free-living PA and physical functioning cross-sectionally and prospectively [19].

The Godin-Shephard Leisure-Time Physical Activity questionnaire was used to measure the frequency of strenuous, moderate, and light leisure-time PA performed for durations of 15 minutes or more over a 7-d period. Participants were asked, *“During a typical 7-d period, how many times on average do you do the following kinds of exercise for more than 15 minutes during your free time?”* The frequency for each intensity category was multiplied by the corresponding metabolic equivalents (METs) of 9, 5, and 3 for strenuous, moderate, and light, respectively. Resulting scores were totaled for a weekly leisure-score index (LSI).

### Statistical Analyses

Normality of distributions and data homogeneity were examined with the Shapiro-Wilk and Levene tests, respectively. Data are shown as means and SD. Non-normally distributed data are shown as medians ± interquartile range (IQR). Between-group differences were evaluated with independent *t*-tests and non-parametric equivalents where appropriate. Substantive differences were determined as a measure of effect size (ES): 0.2 small; 0.5 medium; and 0.8 large. Bivariate correlations were used to examine relationships of interest. The threshold for statistical significance was set *a priori* and defined as two-sided alpha of ≤0.05. Data were analyzed with SPSS (v28; Armonk, NY). Figures were generated using GraphPad Software (v8.3; La Jolla, CA).

## RESULTS

### Participants

Descriptive data are shown in Table 1. Consistent with the study design, there were no between-group differences in age, BMI, or body fat%. According to BMI classifications, 14 of 32 (44%) participants were *“normal weight”* (18.5-24 kg/m^2^), 12 of 32 (37%) were *“overweight”* (25-29.9 kg/m^2^), and 6 of 32 (19%) were *“obese”* (>30 kg/m^2^). Diastolic blood pressure (*p* = 0.031, ES = 0.80) and RPP (*p* = 0.039, ES = 0.75) were significantly higher among SARS-CoV-2 participants.

**Table 1.** Descriptives (*n* = 32).

| Variables | SARS-CoV-2<br>$n = 17$ | Controls<br>$n = 15$ | $p$-value | ES |
| --- | --- | --- | --- | --- |
| Age (y) | 55 ± 11 | 54 ± 14 | 0.814 | |
| Height (m) | 1.64 ± 0.04 | 1.61 ± 0.07 | 0.263 | |
| Mass (kg) | 75 ± 20 | 67 ± 10 | 0.170 | |
| BMI (kg/m<sup>2</sup>) | 27.9 ± 7.0 | 25.9 ± 4.4 | 0.343 | |
| Body Fat (%) | 33.7 ± 7.4 | 32.1 ± 7.7 | 0.554 | |
| HR (bpm) | 71 ± 13 | 66 ± 8 | 0.167 | |
| SBP (mm Hg) | 126 ± 9 | 118 ± 11 | 0.051 | |
| DBP (mm Hg) | 82 ± 9 | 75 ± 7 | 0.031† | 0.78 |
| Rate-pressure product | 89 ± 17 | 77 ± 13 | 0.039† | 0.75 |
| PFSDQ-M [median (IQR)]‡ | 10 (14) | 2 (3) | 0.008† | 0.23 |
| Walking Self-Efficacy [median (IQR)] | 81 (44) | 95 (18) | 0.274 |  |
| Leisure Score Index [median (IQR)] | 21 (18) | 36 (22) | 0.004† | 0.27 |
| Depression Medications (yes) [$n$ (%)] | 6 (35) | 1 (7) | 0.051 | |
| Employed (yes) [$n$ (%)] | 11 (65) | 11 (73) | 0.599 | |
| Marital Status [$n$ (%)] | | | | |
| Married or living with sig. other | 11 (65) | 11 (73) | 0.599 |  |
| Education |  |  |  |  |
| Bachelor's Degree (yes) [$n$ (%)] | 11 (65) | 13 (87) | 0.152 | |
| Annual Income [$n$ (%)] | | | | |
| ≥ \$50k | 13 (76) | 12 (80) | 0.810 | |

Values are shown as means and SD unless noted otherwise. ES, effect size; BMI, body mass index; HR, heart rate; bpm, beats per min; SBP, systolic blood pressure; DBP, diastolic blood pressure; MAP, mean arterial blood pressure; mm Hg, millimeters per mercury; Rate-pressure product calculated by dividing the product of HR and SBP by 100. IQR, interquartile range. PFSDQ-M, Pulmonary Functional Status and Dyspnea Questionnaire. ‡higher number indicative of greater disturbance. †Statistically significant between-group difference at $P$ -value $\leq 0.050$ .

### Symptom Frequency and Duration for SARS-CoV-2 Participants

SARS-CoV-2 symptom frequency and duration are shown in Figure 1. Following a positive diagnosis, median time to study enrollment was 85 (IQR: 61) days. Loss of taste/smell, fever/cough, and joint/muscle aches were the most persistent symptoms affecting 71%, 53%, and 41% of participants, respectively. Though fever and sore throat were common – these symptoms tended to be short-lived – fever lasted 1 to 16 days whereas sore throat lasted 2 to 10 days. Of note, 9 of 17 (53%) SARS-CoV-2 participants were *symptomatic* during testing with a median time since diagnosis of 74 (IQR: 67) days.

**Figure 1.**
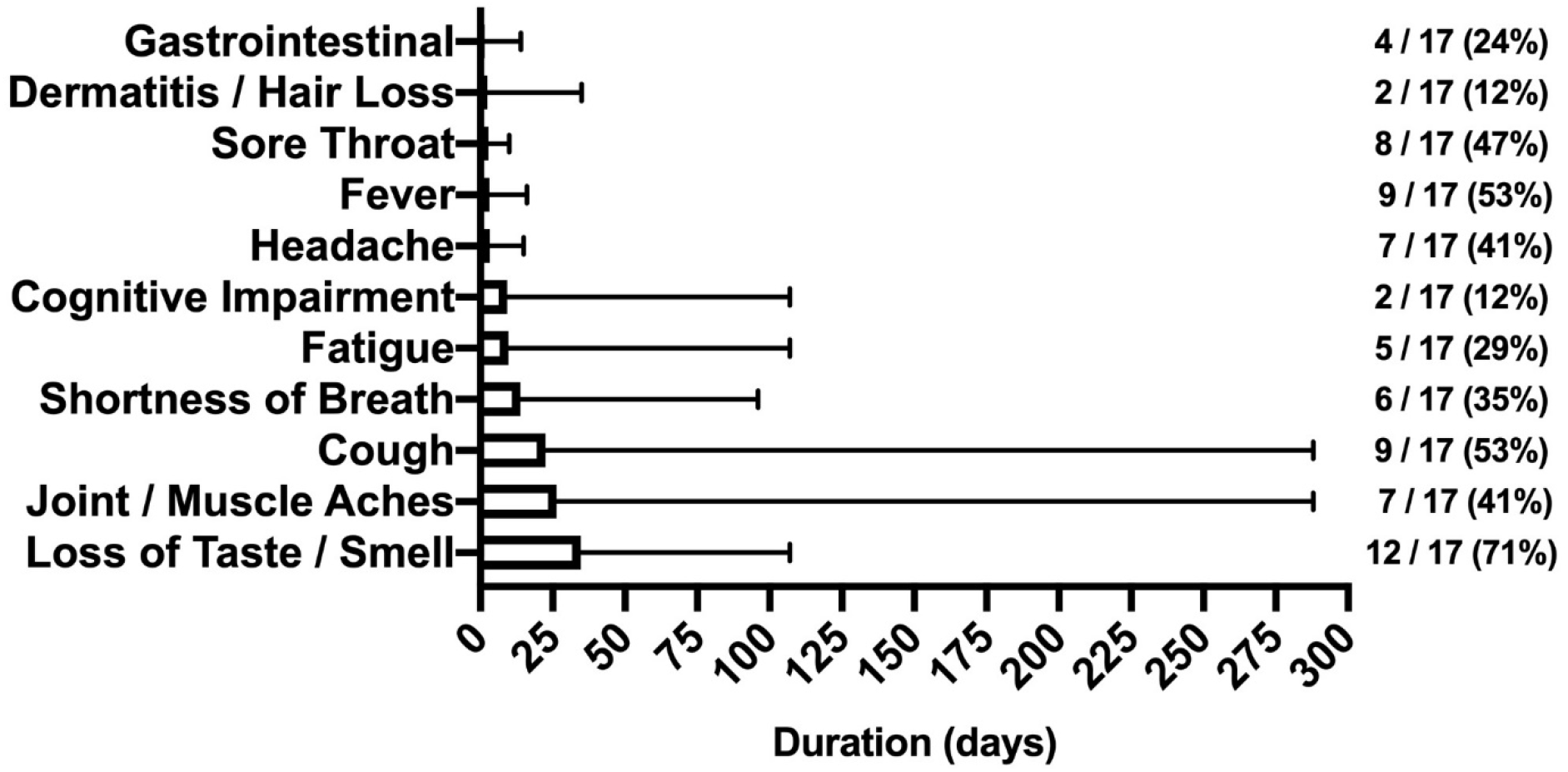
Symptoms reported by SARS-CoV-2 participants. The duration (days) and frequency *n* (%) of symptoms reported after illness onset. Note the protracted duration in cough and joint / muscle aches.

### Between-Group Differences for Functional Status, POMS, and LSI

Though no between-group differences were observed in walking self-efficacy, functional status (*p* = 0.008, ES = 0.23) and LSI (*p* = 0.004, ES = 0.27) were significantly poorer in SARS-CoV-2 participants (Table 1). Significant between-group differences were also detected in POMS TMD (Figure 2) – indicating greater mood disturbance (i.e., more negative mood) in SARS-CoV-2 participants. Three of the six subscales showed SARS-CoV-2 participants had significantly: elevated tension, confusion, and lower vigor (Table 2 and Figure 3).

**Figure 2.**
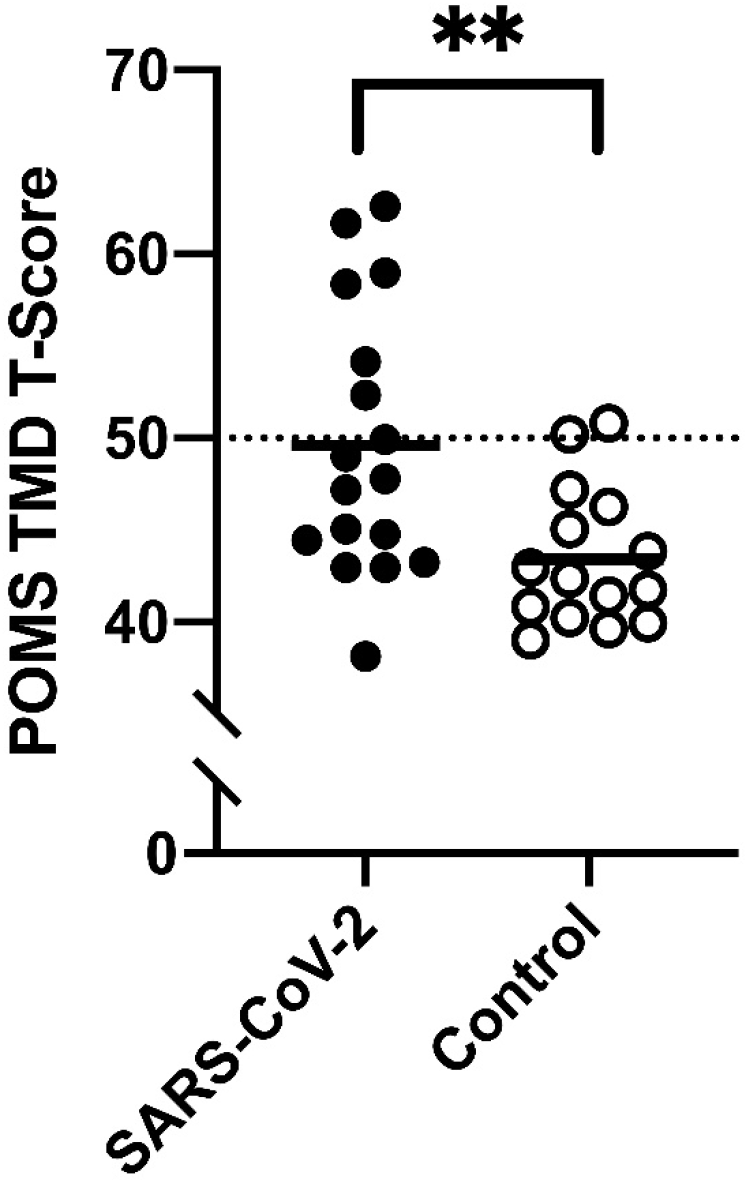
Between-group differences in Total Mood Disturbance (TMD) T-scores from the Profile of Mood States (POMS). \*\**p* < 0.01. Means and 95% confidence intervals: SARS-CoV-2, 49.6 (46.2 to 53.1) *vs*. controls, 43.4 (41.7 to 45.5).

**Table 2.** Profile of Mood States Raw Scores (*n* = 32)

| POMS Score | SARS-CoV-2<br>$n = 17$ | Controls<br>$n = 15$ | $p$-value | ES |
| --- | --- | --- | --- | --- |
| TMD | $19 \pm 24$ | $-1 \pm 12$ | 0.005† | 1.0 |
| Tension [median (IQR)] | 7 (12) | 5 (4) | 0.009† | 0.9 |
| Depression [median (IQR)] | 3 (11) | 3 (3) | 0.262 |  |
| Anger | $6 \pm 5$ | $3 \pm 2$ | 0.132 | |
| Vigor | $15 \pm 6$ | $20 \pm 6$ | 0.043† | 0.7 |
| Fatigue | $6 \pm 6$ | $4 \pm 4$ | 0.265 | |
| Confusion [median (IQR)] | 6 (4) | 3 (2) | 0.005† | 1.1 |

Values are shown as means and SD unless noted otherwise. POMS, Profile of Mood States; ES, effect size; TMD, total mood disturbance. IQR, interquartile range. †Statistically significant between-group differences at $p$ -value $\leq 0.05$ .

**Figure 3.**
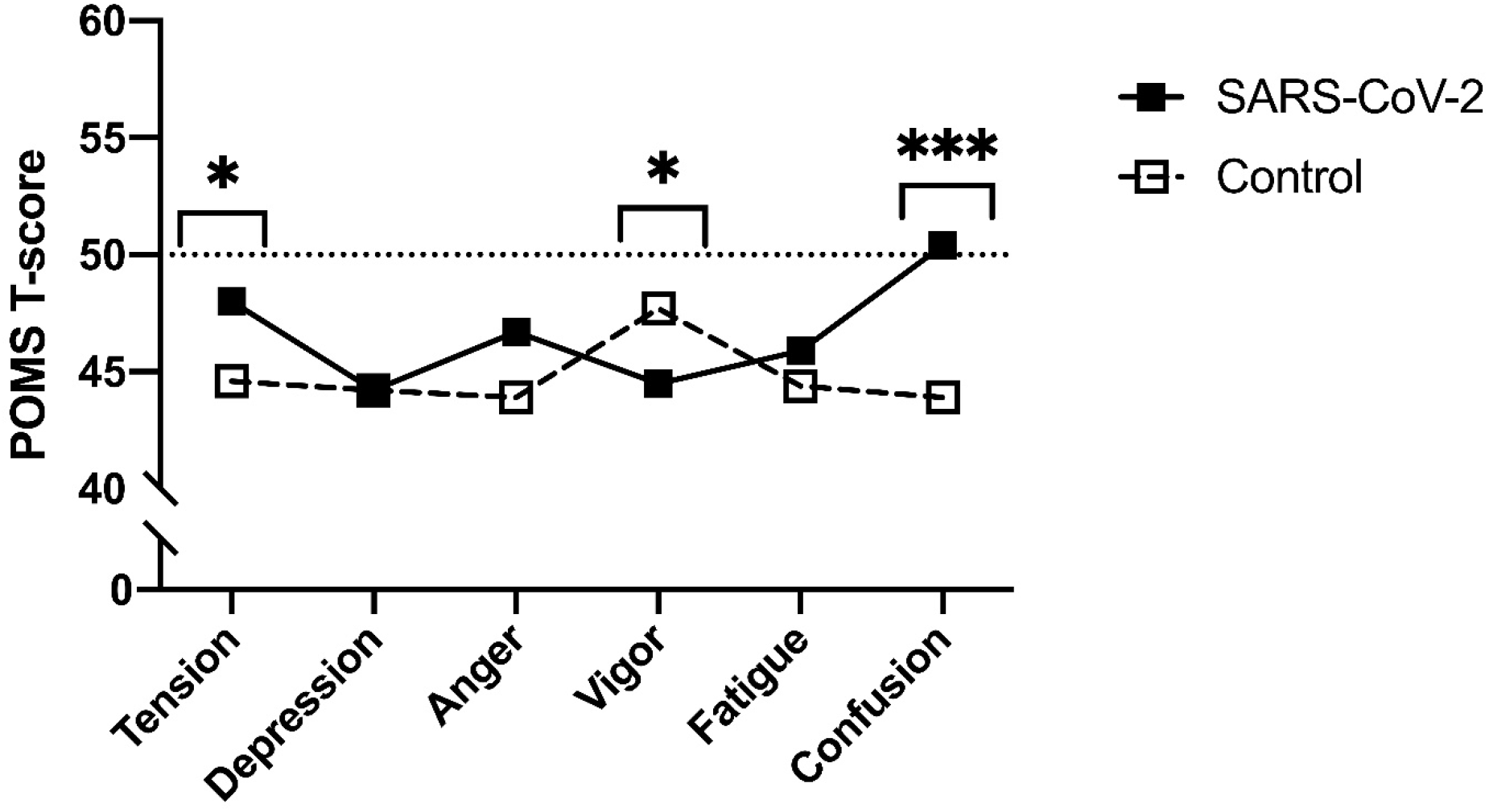
Profile of Mood States (POMS) sub-scale T-scores between SARS-CoV-2 and control participants. \**p* < 0.05. \*\*\**p* < 0.001. Means and 95% confidence intervals. *Tension*: SARS-CoV-2, 52.0 (47.9 to 57.0) *vs*. controls, 44.2 (41.6 to 46.8); *Vigor:* SARS-CoV-2, 44.4 (39.2 to 49.1) *vs*. controls, 51.4 (47.3 to 55.5); *Confusion:* SARS-CoV-2, 51.7 (48.0 to 56.3) *vs*. controls, 44.0 (41.9 to 46.6).

### Relationships between Symptoms, Functional Status, and POMS

The number of symptoms reported at illness onset were positively associated (*r* = 0.620, *p* = 0.008) with confusion (Figure 4A) – interestingly significance persisted, in separate analyses, independent of age (*r* = 0.623, *p* = 0.010) or body fat% (*r* = 0.619, *p* = 0.011). Moreover, functional status was positively associated with the number of days SARS-CoV-2 participants reported loss of taste / smell (Figure 4B). Further examination revealed significant differences between *symptomatic* and *asymptomatic* SARS-CoV-2 participants in walking self-efficacy (median +/- IQR) (asymptomatic; 100 ± 23 *vs*. symptomatic; 60 ± 43, *p* = 0.032, ES = 0.29) and functional status (median +/- IQR) (asymptomatic; 4 ± 9 *vs*. symptomatic; 15 ± 15, *p* = 0.005, ES = 0.49). Functional status was positively associated with fatigue (*r* = 0.491, *p* = 0.045) among SARS-CoV-2 participants whereas functional status was negatively associated with anger (*r* = -0.585, *p* = 0.022) among control participants. In the total sample, functional status and walking self-efficacy were negatively associated (*r* = -0.420, *p* = 0.017). Additionally, RPP was negatively associated with LSI (*r* = -0.605, *p* < 0.001) and walking self-efficacy (*r* = -0.353, *p* = 0.047) among all participants.

**Figure 4.**
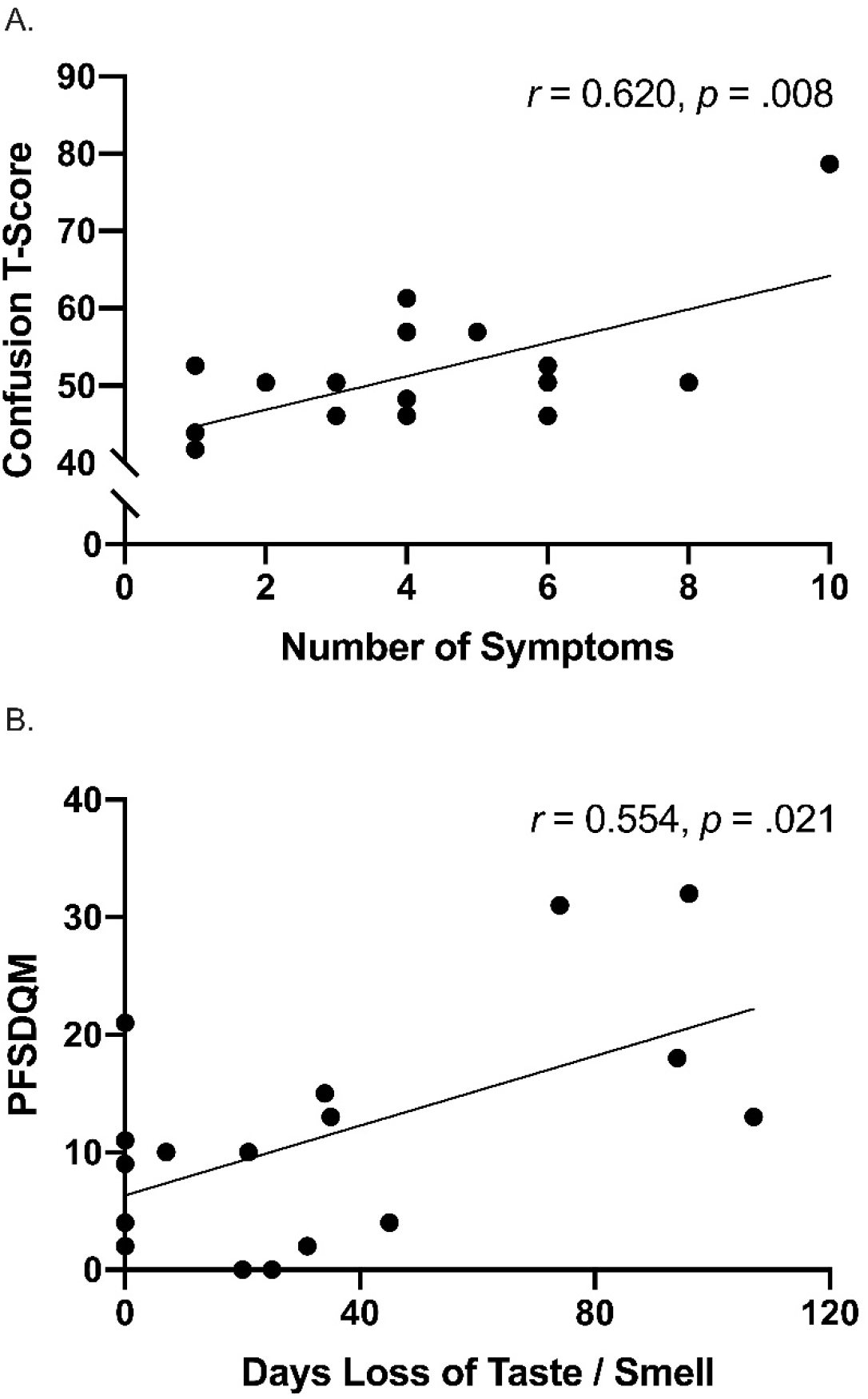
A) Unadjusted scatterplot between number of symptoms reported after illness onset and confusion T-score. B) unadjusted scatterplot between functional status and number of days experiencing loss of taste / smell.

## DISCUSSION

The direct and indirect effects of the COVID-19 pandemic remain a concern to personal and public health. Herein, we compared functional status, mood state, and leisure-time PA between women with and without a history of SARS-CoV-2 matched for age, BMI, smoking status, and history of cardiopulmonary disease. In agreement with our hypotheses, SARS-CoV-2 participants reported greater TMD than controls. With respect to normative data, the control group possessed a lower TMD score (i.e., more positive mood), whereas 6 of 17 (35%) SARS-CoV-2 participants exhibited TMD scores above the normative data (i.e., more negative mood). Significant between-group differences were also found in three of the six POMS sub-scales, in which, the SARS-CoV-2 group reported elevated tension, confusion, and lower vigor. Of note, the confusion scores among SARS-CoV-2 participants were significantly higher than a comparable group of adult women [16]. A significant between-group difference in vigor, but not fatigue was observed, however; research evaluating mood state in response to exercise training suggests fatigue and vigor perform orthogonally rather than as polar endpoints of a single continuum [20].

There is a precedent for mental health disturbance in previous coronavirus epidemics including severe acute respiratory syndrome (SARS) and Middle East respiratory syndrome (MERS) [21]. In these instances, depressed mood and memory impairment were documented during the acute viral infection as well as during varying periods in the post-acute recovery phase among hospitalized individuals. Emerging evidence now suggests SARS-CoV-2 may follow a similar time-course with the number of SARS-CoV-2 symptoms reported at illness onset being linked to the development of clinically significant depression, anxiety, and post-traumatic stress symptoms in recovery [22]. Along these lines, we observed the number of reported initial SARS-CoV-2 symptoms were positively associated with confusion sub-scale scores. However, it remains unclear whether related mental health consequences of SARS-CoV-2 are attributable to the direct or indirect effects of the virus. Some have speculated that retrograde axonal transport from peripheral nerves including the gustatory and olfactory nerves may be a route to central nervous system infiltration [23]. In the present work, 12 of 17 (71%) SARS-CoV-2 participants reported loss of taste / smell which corresponded with increased tension (*r* = 0.533, *p* = 0.028) and depression (*r* = 0.442, *p* = 0.076) – providing plausibility for this speculation.

To date, limited investigations have evaluated physical functioning in SARS-CoV-2 survivors – particularly among those with post-acute COVID-19 syndrome. It is known that impaired physical function, owning to the effects of dyspnea, fatigue and joint / muscle pain, anxiety, and depression can interfere with the ability to independently perform activities of daily living. Regrettably, this can extend into the realm of personal care such that limited mobility can degrade quality-of-life outcomes. A recent systematic review found that functional ability, as assessed by questionnaire, appears compromised during the acute phase of SARS-CoV-2 [24]. These acute effects are expected given the well-documented respiratory and cardiovascular manifestations of the virus. However, in the present work, post-acute SARS-CoV-2 participants had poorer functional status and reported less leisure-time PA compared to controls – despite a median time of 74 days since diagnosis. Notably, functional status (where a higher number corresponds to more disturbance) was positively associated with fatigue among SARS-CoV-2 participants. A possible interpretation of these interrelated factors is that increased feelings of fatigue may complicate activities of daily living, or alternatively, activities of daily living may overburden symptom recovery thereby contributing to increased feelings of fatigue. Hence, periodic assessment of functional status using validated questionnaires such as the PFSQD-M may offer relevant information about the recovery time-course and impact on psychosocial health and quality-of-life in post-acute COVID-19 syndrome.

While varying circumstances including preexisting comorbidities can influence the path to recover, there is evidence suggesting women may be disproportionately affected by post-acute COVID-19 syndrome [26,27]. To an extent, it would be expected that advancing age and excess body weight are also risk factors for delayed recovery as occult and/or overt disease are known to increase vulnerability to disease [8]. Sudre and colleagues [27] recently showed that experiencing five or more symptoms during the initial week of illness was associated with an increased risk of post-acute COVID-19 syndrome. Interestingly, in the present work, SARS-CoV-2 participants who were symptomatic at the time of testing reported 5 ± 3 symptoms at the time of illness onset. These participants were also found to have lower walking self-efficacy and poorer functional status independent of age or BMI (compared to asymptomatic SARS-CoV-2 participants). Thus, it seems functional impairment accompanying post-acute COVID-19 syndrome may be worse in individuals wherein the SARS-CoV-2 virus elicited more symptoms reported during acute illness.

It is understandable that persistent SARS-CoV-2 symptoms are likely to degrade functional status while also inciting feelings of concern about health and well-being. Indeed, leisure-time PA was negatively associated (*r* = -0.672, *p* = 0.047) with the number of symptoms among symptomatic SARS-CoV-2 participants in the present work. This finding is consistent with process of interoception wherein peripheral (sensory) feedback provides a moment-to-moment updates of the internal bodily environment [28]. This, in turn, has the potential to trigger a maladaptive cycle that can ultimately interfere with activities of daily living. Given that some SARS-CoV-2 symptoms, notably cough and joint/muscle aches can last beyond 3-4 months, systemic deconditioning may become a lasting complication. This becomes especially important considering women already tend to exhibit greater rates of age-related disability than men [7] – and female sex is a risk factor for persistent SARS-CoV-2 symptoms [27]. Within this context, further work is needed to develop and implement targeted interventions to offset the burden of deconditioning in post-acute COVID-19 syndrome.

### Limitations

To our knowledge, no prior work has examined constructs of mental and physical health in SARS-CoV-2 while employing a case-control design matching participants on age, BMI, smoking status, and history of cardiopulmonary disease. As such, we have reasonable confidence that our observations are due to meaningful between-group differences in functional status, mood state, and leisure-time PA. Nevertheless, there are several limitations that need to be considered. First, the retrospective nature of the work limits our ability to know whether alterations in the observed outcomes were present before testing. Second, because the study sample was purposely limited to women, findings may not be generalizable to men. Third, since antibody testing was not performed, it is unknown if control participants might have unknowingly had a prior SARS-CoV-2 infection. Lastly, we acknowledge a relatively small sample size. However, given the strength of the reported effect sizes – we reached statistical significance across multiple measures. Future work should examine prospective, longitudinal changes in functional status, mood state, and leisure-PA in post-acute COVID-19 syndrome while taking into consideration the related impact on quality-of-life and objective measures of PA. Such information will prove useful in our collective understanding about the sequela of persistent symptoms that can undermine mental and physical health in post-acute COVID-19 syndrome.

### Conclusions

The present work extends the growing evidence highlighting the adverse consequences of post-acute COVID-19 syndrome on mental health and functional status in women. We report greater TMD – attributed to increased tension, increased confusion, and decreased vigor in SARS-CoV-2 participants. Likewise, poorer functional status was noted which was positively associated with fatigue among SARS-CoV-2 participants. Interestingly, among SARS-CoV-2 participants, the number of symptoms reported at illness onset were positively associated with confusion.

## Data Availability

All data produced in the present study are available upon reasonable request to the authors

## Acknowledgements

The authors greatly appreciate the recruitment efforts of the Indiana Clinical and Translational Sciences Institute’s All IN for Health team and Tyler B. Baker for his assistance with data collection.

## Author Contributions

All testing was performed in the Human Performance Laboratory at the Indiana University School of Public Health – Bloomington. SJC and MNB conceptualized the study design. SJC obtained funding, provided supervision of experimental testing, performed data analyses, and drafted the original manuscript. MNB conducted experimental testing, completed data analyses, generated figures, and provided revisions to manuscript drafts. JSR, BAP, and BLP gave guidance, contributed intellectual content, and provided revisions to manuscript drafts. All authors approved of the final version of the manuscript.

## Notes

The authors have no perceived or potential conflicts of interest, financial or otherwise to declare.

### Competing Interest Statement

The authors have declared no competing interest.

### Funding Statement

This study was funded by the Indiana Clinical and Translational Sciences Institute by grant number UL1TR002529 from the NIH National Center for Advancing Translational Sciences, Clinical and Translational Sciences Award.

### Author Declarations

Ethics committee/IRB of Indiana University gave ethical approval for this work.

